# Mortality among healthcare workers in Indonesia during 18 months of COVID-19

**DOI:** 10.1101/2022.04.27.22274334

**Authors:** Lenny L. Ekawati, Ahmad Arif, Irma Hidayana, Ahmad Nurhasim, M. Zakiyuddin Munziri, Karina D. Lestari, Amanda Tan, Firdaus Ferdiansyah, Fikry Nashiruddin, Qorinah E.S. Adnani, Halik Malik, Tri Maharani, Andy Riza, Monalisa Pasaribu, Khairul Abidin, Adhi A. Andrianto, Nursalam, A.V. Sri Suhardiningsih, Ade Jubaedah, N.S. Widodo, Henry Surendra, Herawati Sudoyo, Adrian D. Smith, Philip Kreager, J. Kevin Baird, Iqbal R.F. Elyazar

**Affiliations:** Eijkman-Oxford Clinical Research Unit (EOCRU), Jakarta, Indonesia; Nuffield Department of Medicine, University of Oxford, United Kingdom; LaporCOVID-19, Jakarta, Indonesia; St. Lawrence University, U.S.A.; The Conversation, Jakarta, Indonesia; Faculty of Medicine, Padjadjaran University, Bandung, Indonesia; Indonesian Doctor Association, Jakarta, Indonesia; National Institute of Health Research and Development, Ministry of Health, the Republic of Indonesia, Jakarta, Indonesia; Indonesia National Nurses Association of East Java Province, Surabaya, Indonesia; Faculty of Nursing, Airlangga University, Surabaya, Indonesia; Indonesian Midwives Association, Jakarta, Indonesia; Association of Indonesian Medical Laboratory Technologist, Jakarta, Indonesia; Mochtar Riady Institute for Nanotechnology, Banten, Indonesia; Nuffield Department of Population Health, University of Oxford, Oxford, United Kingdom; Institute of Human Sciences, University of Oxford, Oxford, United Kingdom

**Keywords:** COVID-19, mortality, healthcare workers, resilience, digital cemetery

## Abstract

The impact of SARS-CoV-2 infections upon Indonesian health care workers (HCWs) remains unclear, as mortality data specific to HCWs is not systematically collected or analyzed in this setting. This report describes findings from a systematic collation, abstraction and analysis of HCW fatalities during the first 18 months of COVID-19 in Indonesia. HCW who died during the period of March 2020 to July 2021 across Indonesia were identified on *Pusara Digital*, a community web-based digital cemetery database dedicated to HCW. We calculated mortality rates and death risk ratio among HCWs and the general population. Qualitative methods explored concerns regarding mortality among HCWs. The analysis suggests that at least 1,545 HCWs died during the study period. The death of males and females HCWs were almost equally distributed (51% vs. 49%). Most were medical doctors and specialists (535, 35%), nurses (428, 28%), and midwives (359, 23%). Deaths most frequently occurred in the age group of 40 to 59 years old with the median age of 50 years (IQR: 39-59). At least 322 (21%) deaths occurred with pre-existing conditions, including 45 who were pregnant. We estimated a minimal HCW mortality rate in Indonesia at 1.707 deaths per 1000 HCW during the first 18 months of COVID-19. Provincial HCW mortality rates ranged from 0.136 (West Sulawesi) to 5.32 HCW deaths per 1000 HCWs (East Java). HCW had a significantly higher mortality rate than the general population (RR = 4.92, 95% CI 4.67 – 5.17). The COVID-19 event in Indonesia resulted in the loss of many hundreds of HCWs, most of them being senior physicians, nurses, and midwives. The HCW death rates is 5-times higher than everyone else. The sheer sparseness of the workforce requires more protective steps and a national systematic surveillance of occupational mortality is urgently needed in this setting.

## Introduction

The disease resulting from nearly 500 million confirmed SARS-CoV-2 infections (COVID-19) had caused approximately 6 million deaths globally [1]. Healthcare workers (HCWs) serving these patients are at high risk of becoming infected [2, 3]. In the early months of the COVID-19 pandemic, a survey of morbidity and mortality on HCWs in 130 countries counted hundreds of deaths [4]. After 18 months, among 135 million HCWs, World Health Organization (WHO) estimated at least 115,000 (range: 80,000 – 180,000) HCWs died as a consequence of SARS-CoV-2 infection [5].

The first cluster of SARS-CoV-2 infections in Indonesia was detected in early March 2020 [6]. Later in mid-2021, the Delta variant of SARS-CoV-2 dominated the surge [7, 8], and overwhelmed hospital capacities across Indonesia [8]. The surge in Delta variant cases prompted stricter emergency restrictions on people’s movement and gathering [9]. SARS-CoV-2 test positivity rates during this crisis period approached 40% [9]. The rapid rise of hospitalizations, emergency care units, scarcity of supplies, and ICU bed needs outstripping availability have caused HCW anxiety and exhaustion [10].

The high proportion of COVID-19 cases among HCWs working in patient ward was identified [11, 12], and their risk of contracting COVID-19 is higher than the general population [13]. Logic evidence informs safe standard operating procedures (SOPs) in healthcare settings [14], personal protective equipment (PPE) inspection, and HCWs practice review are critically important to avoid negligence in caring for infectious COVID-19 patients [15-17]. Inadequate PPE, or being unfamiliar with wearing it has contributed to a substantially elevated risk of COVID-19 compared to other groups not working in a hospital setting [18-20]. Working environment, type of occupation, length of exposure with COVID-19 patients, and testing availability have been identified as factors associated with SARS-CoV-2 infection among HCWs [20-23].

Heavy burden in managing COVID-19 patients may lead to emotional exhaustion and jeopardize professional efficacy [24]. However, a manageable working schedule, mental health, psychosocial support, remuneration and incentives are required for HCWs’ welfare [25-27]. Safety issues and high protective working conditions for pregnant HCWs and those with higher vulnerability due to age, ethnicity, social determinants, or underlying conditions should be implemented and carefully monitored [28].

The Indonesian Digital Cemetery (*Pusara Digital;* https://nakes.laporcovid19.org/) by far is the most comprehensive source of data on HCW mortality in Indonesia. *Pusara Digital* received and recorded mortality data as both public service and memorial to those lives lost in humanitarian service. The study represents an effort to utilize that resource and provides a descriptive analysis of mortality due to COVID-19 among HCWs during 2020-2021. This report derives a systematic quantitative estimate of COVID-19 mortality among HCWs in Indonesia, and its comparison with general population. We qualitatively explore family, friends, and colleagues’ concerns regarding their loss.

## Methods

### Source of routine information

The daily verified COVID-19 confirmed cases and death data from 34 provinces were available on the government website, managed by the Center of Health Data of the Indonesian Ministry of Health (MoH) [29]. The research team extracted the daily provincial-level aggregated data recorded between March 2020 and July 2021, then entered the extracted data into an Excel database. The number of population and the number of HCW by provinces were extracted from the 2020 Indonesia Annual Health Profile [30].

### Collection of demographic and health information for HCW deaths

The research team downloaded HCW mortality data from Pusara Digital, a web-based digital cemetery database dedicated to HCW’s. The platform is operated by LaporCOVID-19, a registered not-for-profit organization established in March 2020, by public health experts, scientists, journalists, and social activists committed to the evidence-based and unbiased characterization of the social economic, and health impacts of COVID-19 on the Indonesian people.

LaporCOVID-19 organized 35 volunteers to manage *Pusara Digital* website in September 2020 and has undertaken two methods of HCW mortality data searching. First, the volunteers looked for daily obituaries on online news and social media (Twitter, Facebook, Instagram). Second, they received data from professional medical and health associations: Association of Indonesian Medical Doctors (*Ikatan Dokter Indonesia; IDI*); National Association of Nurses (*Persatuan Perawat Nasional Indonesia, PPNI*); Association of Midwives (*Ikatan Bidan Indonesia, IBI*); and Association of Medical Laboratory Technologists (*Persatuan Ahli Teknologi Laboratorium Kesehatan Indonesia, PATELKI*). From those sources, the volunteers curated essential variables and organized testimonials from surviving family and colleagues that were sent to the website.

Data were entered into an Excel database with limited access and password protected facility. The variables consisted of full name, title, age, gender, education, healthcare work type, work setting, latitude and longitude of district/city, underlying health condition, date of birth and death, and COVID-19 diagnostic status (suspect, probable, or confirmed) defined by the Indonesian MoH [31]. All individual information of the deceased is treated as confidential. After January 2021, when the government of Indonesia rolled out the COVID-19 vaccination, the team sought and recorded the official vaccination status of each deceased (dates and type of vaccination by, and vaccine’s name). HCWs were afforded top priority in that roll out scheme [32].

### Assembling community narratives relating to HCW death

*Pusara Digital* generates obituaries on the website along with dedicated space to enable testimonial submissions. This feature allows families, friends, and communities to write testimonials and commemorate HCWs under their obituary and in the front page of the website. Submitted testimonials from the community, colleagues, and family of the deceased serve as an open-source data for verification, corrections, and revisions, which enable us to verify the information on the website. The system records name, phone number, submission date and acceptance, content, and the designated HCWs. The team extracted over 2,600 records of unstructured notes and comments from September 2020 to July 2021.

### Statistical analysis

HCW mortality frequency was cross-tabulated by geography, gender, age, occupational type, working place, COVID-19 status, COVID-19 patient care, and vaccination status. We calculated mortality rate of the general population by dividing the number of reported deaths by the population at risk during pandemic. HCW mortality rate was calculated by dividing the number of HCW reported deaths and the number of HCW at risk during pandemic. The number of population and the number of HCW by provinces were extracted from the 2020 Indonesia Annual Health Profile [30].

We computed the death risk ratio to quantify the likelihood of the death for HCW group against the risk of death in the general population. A 95% confidence interval of risk ratio was calculated using the log-transformation method. Mortality rate and risk ratio analysis was carried out at provincial, island, and national level over time. The statistical correlation between HCW mortality rate and the general population mortality rate was calculated using Spearman’s rank correlation and p-value = 0.05 as a threshold of statistical significance. Data were analyzed using Stata statistical version 12 (StataCorp, College Station, TX, USA). Mapping of provincial mortality rates used QGIS mapping software version 3.16.6 [33].

### Qualitative analysis

Content analysis was applied to qualitatively explore people’s concerns regarding HCW mortality [34, 35]. This method quantitatively summarizes the narrative responses from key categories [36-38], such as groups of senders (families, colleagues, former patients, teachers/students, or community) and content of the testimonials (personals, professionals or both). The concept unit of analysis refers to the submitted written texts, which were divided into smaller units and coded [39, 40].

The method of Graneheim and Lundman [40] was applied for validation and consistency. Further, a manifest analysis was performed to describe what the commentators mentioned regarding the loss of HCWs’ life. This type of analysis solely relies on the words to illustrate the visible and obvious meaning of the messages [38, 39]. Content analysis allows any number of informants or written text to be explored [41]. Coding and analysis were conducted by using NVivo version 12 (QSR International Pty Ltd, released in 2018).

### Ethical approval

This study was approved by the Health Research Ethics Committee of the National Institute of Health Research and Development, Ministry of Health, Republic of Indonesia (LB.02.01/2/KE/620/2020).

## Results

### HCWs mortality

*Pusara Digital* documented 1,545 known HCW deaths during the first 18 months of COVID-19 pandemics (Table 1). The deaths between males and females slightly differed (51% vs 49%). The median age among lost HCWs was 50 years (IQR: 39-59 years). The largest age group was 50– 59-year-olds (287 individuals, 19%), followed by HCWs less than 40 years old, 40–49-year-olds (223 individuals, 14%), and 30-39 years old (185 individuals, 12%).

**Table 1.** Individual characteristics of deceased healthcare workers between March 2020 and July 2021 in Indonesia.

| Characteristics | Sumatra | Java | Bali & Nusa Tenggara | Kalimantan | Sulawesi | Maluku & Papua | Total | Percent |
| --- | --- | --- | --- | --- | --- | --- | --- | --- |
| **Total HCW population<sup>1</sup>** | **232116** | **418970** | **59207** | **65778** | **96992** | **32198** | **905261** | **100%** |
| **Total HCW deaths<sup>2</sup>** | **163** | **1208** | **29** | **68** | **63** | **14** | **1545** | **0.2%** |
| **Mortality rate per 1,000** | **0.702** | **2.883** | **0.490** | **1.034** | **0.650** | **0.435** | **1.707** |  |
| **Gender** |  |  |  |  |  |  |  |  |
| Male | 86 | 607 | 8 | 43 | 36 | 5 | 785 | 50.8% |
| Female | 77 | 601 | 21 | 25 | 27 | 9 | 760 | 49.2% |
| **Age group (years)** |  |  |  |  |  |  |  |  |
| 20 - 29 | 9 | 52 | 4 | 1 | 4 | 0 | 70 | 4.5% |
| 30 - 39 | 14 | 152 | 4 | 7 | 6 | 2 | 185 | 11.9% |
| 40 - 49 | 32 | 167 | 5 | 9 | 8 | 2 | 223 | 14.4% |
| 50 - 59 | 33 | 223 | 2 | 13 | 12 | 4 | 287 | 18.7% |
| 60 - 69 | 18 | 100 | 2 | 5 | 10 | 2 | 136 | 8.9% |
| 70+ | 6 | 79 | 3 | 3 | 7 | 0 | 98 | 6.3% |
| Missing data | 51 | 435 | 9 | 30 | 16 | 4 | 545 | 35.3% |
| **Age (median, IQR)** | 50<br>(42-56) | 50<br>(39-59) | 46<br>(32-59) | 50<br>(42-58) | 52<br>(43-65) | 51<br>(45-55) | 50<br>(39-59) |  |
| **Occupational type** |  |  |  |  |  |  |  |  |
| Nurses | 26 | 362 | 6 | 21 | 9 | 3 | 427 | 27.6% |
| Midwives | 44 | 283 | 10 | 7 | 11 | 4 | 359 | 23.2% |
| General Practitioners | 30 | 228 | 6 | 12 | 17 | 1 | 294 | 19.0% |
| Specialists | 46 | 166 | 2 | 12 | 13 | 2 | 241 | 15.6% |
| Pharmacists | 5 | 38 | 1 | 5 | 2 | 2 | 53 | 3.2% |
| Laboratory Technologists | 4 | 35 | 4 | 2 | 2 | 2 | 49 | 3.2% |
| Public Health Officers | 2 | 27 | 0 | 5 | 5 | 0 | 39 | 2.5% |
| Dentists | 0 | 31 | 0 | 2 | 3 | 0 | 36 | 2.3% |
| Administrative Staff | 2 | 8 | 0 | 0 | 0 | 0 | 10 | 0.6% |
| Radiographers | 1 | 8 | 0 | 1 | 0 | 0 | 10 | 0.6% |
| Nutritionists | 1 | 7 | 0 | 0 | 1 | 0 | 9 | 0.6% |
| Others | 2 | 15 | 0 | 1 | 0 | 0 | 18 | 1.2% |
| **COVID-19 status** |  |  |  |  |  |  |  |  |
| Positive | 149 | 1137 | 26 | 61 | 59 | 11 | 1443 | 93.4% |
| Probable | 14 | 71 | 3 | 7 | 4 | 3 | 102 | 6.6% |
| **Working place** |  |  |  |  |  |  |  |  |
| Hospital | 68 | 523 | 12 | 34 | 28 | 5 | 670 | 43.4% |
| Primary Health Center | 41 | 264 | 4 | 18 | 12 | 5 | 344 | 22.3% |
| Clinical/Private Practice | 14 | 147 | 3 | 1 | 5 | 0 | 170 | 11.0% |
| Health and Medical School | 9 | 55 | 3 | 1 | 6 | 1 | 75 | 4.9% |
| Other | 31 | 219 | 7 | 14 | 12 | 3 | 286 | 18.5% |
| **COVID-19 patient care** |  |  |  |  |  |  |  |  |
| Yes | 72 | 484 | 7 | 29 | 21 | 3 | 616 | 39.9% |
| Probable | 38 | 345 | 8 | 14 | 17 | 5 | 427 | 27.6% |
| No | 53 | 379 | 14 | 25 | 25 | 6 | 502 | 32.5% |
| **Period of death and vaccination status** |  |  |  |  |  |  |  |  |
| Pre-vaccination campaign | 90 | 499 | 15 | 41 | 35 | 6 | 686 | 44.4% |
| During vaccination campaign | 1 | 3 | 0 | 0 | 1 | 1 | 5 | 0.3% |
| Post-vaccination campaign |  |  |  |  |  |  |  |  |
| - Vaccinated | 4 | 28 | 0 | 0 | 2 | 0 | 34 | 2.2% |
| - Unvaccinated | 10 | 56 | 4 | 4 | 1 | 2 | 77 | 5.0% |
| - Unavailable vaccination status | 58 | 622 | 10 | 22 | 25 | 6 | 743 | 48.1% |
| **Comorbid** |  |  |  |  |  |  |  |  |
| Yes | 24 | 218 | 4 | 12 | 12 | 7 | 277 | 17.9% |
| No | 139 | 990 | 25 | 56 | 51 | 7 | 1268 | 82.1% |
| **Total comorbid** |  |  |  |  |  |  |  |  |
| 1 | 18 | 144 | 3 | 10 | 9 | 5 | 189 | 68.2% |
| 2 | 6 | 54 | 1 | 1 | 3 | 2 | 67 | 24.2% |
| ≥3 | 0 | 20 | 0 | 1 | 0 | 0 | 21 | 7.6% |
| **Comorbid type** |  |  |  |  |  |  |  |  |
| Asthma | 0 | 16 | 0 | 1 | 0 | 0 | 17 | 6.1% |
| Autoimmune disorder | 0 | 3 | 0 | 1 | 0 | 0 | 4 | 1.4% |
| Cancer | 2 | 5 | 0 | 0 | 0 | 1 | 8 | 2.9% |
| Cardiovascular disease | 2 | 22 | 0 | 1 | 0 | 1 | 26 | 9.4% |
| Dengue fever | 0 | 2 | 0 | 0 | 0 | 0 | 2 | 0.7% |
| Diabetes mellitus | 9 | 104 | 1 | 3 | 5 | 2 | 124 | 44.8% |
| Gastrointestinal disease | 1 | 1 | 0 | 0 | 0 | 0 | 2 | 0.7% |
| Haematological disorder | 0 | 1 | 0 | 0 | 0 | 0 | 1 | 0.4% |
| Hepatitis | 0 | 2 | 0 | 0 | 0 | 0 | 2 | 0.7% |
| Hernia nucleus pulposus | 0 | 1 | 0 | 0 | 0 | 0 | 1 | 0.4% |
| HIV | 0 | 0 | 0 | 0 | 1 | 0 | 1 | 0.4% |
| Hypertension | 3 | 22 | 0 | 3 | 1 | 2 | 31 | 11.2% |
| Hyperthyroid | 0 | 2 | 0 | 0 | 0 | 0 | 2 | 0.7% |
| Kidney disease | 0 | 4 | 0 | 0 | 1 | 0 | 5 | 1.8% |
| Neurological disorder | 0 | 1 | 0 | 0 | 0 | 0 | 1 | 0.4% |
| Obesity | 0 | 7 | 0 | 0 | 1 | 1 | 9 | 3.2% |
| Parkinson disease | 0 | 1 | 0 | 0 | 0 | 0 | 1 | 0.4% |
| Pneumonia | 0 | 2 | 1 | 0 | 0 | 0 | 3 | 1.1% |
| Pre-eclampsia | 1 | 0 | 0 | 0 | 0 | 0 | 1 | 0.4% |
| Respiratory disease | 1 | 1 | 0 | 0 | 0 | 0 | 2 | 0.7% |
| Stroke | 0 | 1 | 0 | 0 | 0 | 0 | 1 | 0.4% |
| Thyroid | 1 | 0 | 0 | 0 | 0 | 0 | 1 | 0.4% |
| Typhus | 1 | 1 | 0 | 0 | 0 | 0 | 2 | 0.7% |
| Unknown | 3 | 19 | 2 | 3 | 3 | 0 | 30 | 10.8% |

Data sources:
<sup>1</sup>The Indonesian Ministry of Health, as per December 31, 2020, http://bppsdmk.kemkes.go.id/info_sdmk/info/#
<sup>2</sup>Pusara Digital LaporCOVID-19, as per July 31, 2021, https://nakes.laporcovid19.org/

Among 1,545 HCW deaths, 535 (35%) were general practitioners and specialized medical practitioners, 428 (28%) nurses, 359 (23%) midwives, 50 (3%) pharmacists, 49 (3%) medical laboratory technologists, 36 (2%) dentists and others (Table 1). There were 1,443 (93%) HCWs deaths with confirmed positive COVID-19, while the remaining 102 (7%) were probable COVID-19 deaths. Underlying health conditions were recorded in 277 (18%) HCWs, including diabetes mellitus (45%), hypertension (11%), cardiac disease (9%), asthma (6%), cancers (3%), and obesity (3%). Among those with comorbidities, just one occurred among 189 (62%) deceased of HCWs, while two combinations of comorbid, and more were revealed from 67 (24%) and 21 (8%) HCWs, respectively.

Forty-five female HCWs (5.9%) were known to have been pregnant prior to death. Sixteen delivered preterm infants immediately prior to death, and 29 died while pregnant. Among them, nine (18%) died before the vaccination program was launched, while 35 (78%) HCWs were not received the vaccine [42] (range time of death: March 19 to July 30, 2021). Only one pregnant HCW who died in May 2021 had received her first dose.

All settings of healthcare delivery were represented in this study: hospitals (670, 43%), primary health centers (344, 22%), private practices and health clinics (170, 11%), health and medical school (75, 5%), and others (286, 19%). Approximately 39.9% of the deceased were directly involved in routine COVID-19 patient management, whereas over half of them may have been occasionally on the COVID-19 wards (27.6%) or were not assigned (32.5%) in the COVID-19 ward.

The first HCW deaths were recorded on 12^th^ March 2020, just 10 days after Indonesia’s first confirmed COVID-19 case cluster was detected. Figure 1C illustrates the number of daily deaths among HCWs. The highest daily death occurred on December 18, 2020 (11) and January 6, 2021 (10), with the average death per day was five HCWs (range: 0-11 individuals). Starting the last days of February 2021, the number of late HCW steeply plummeted and continued to decline until May 2021 with a total of 25 deaths. Nonetheless, dramatic leaps occurred from mid-June to July 2021, when the highest daily fallen HCWs accounted for 14 individuals on June 24, 2021 and 26 individuals on July 8, 2021. The average death per day was 10 HCWs, ranging from 0-26 individuals.

**Figure 1.**
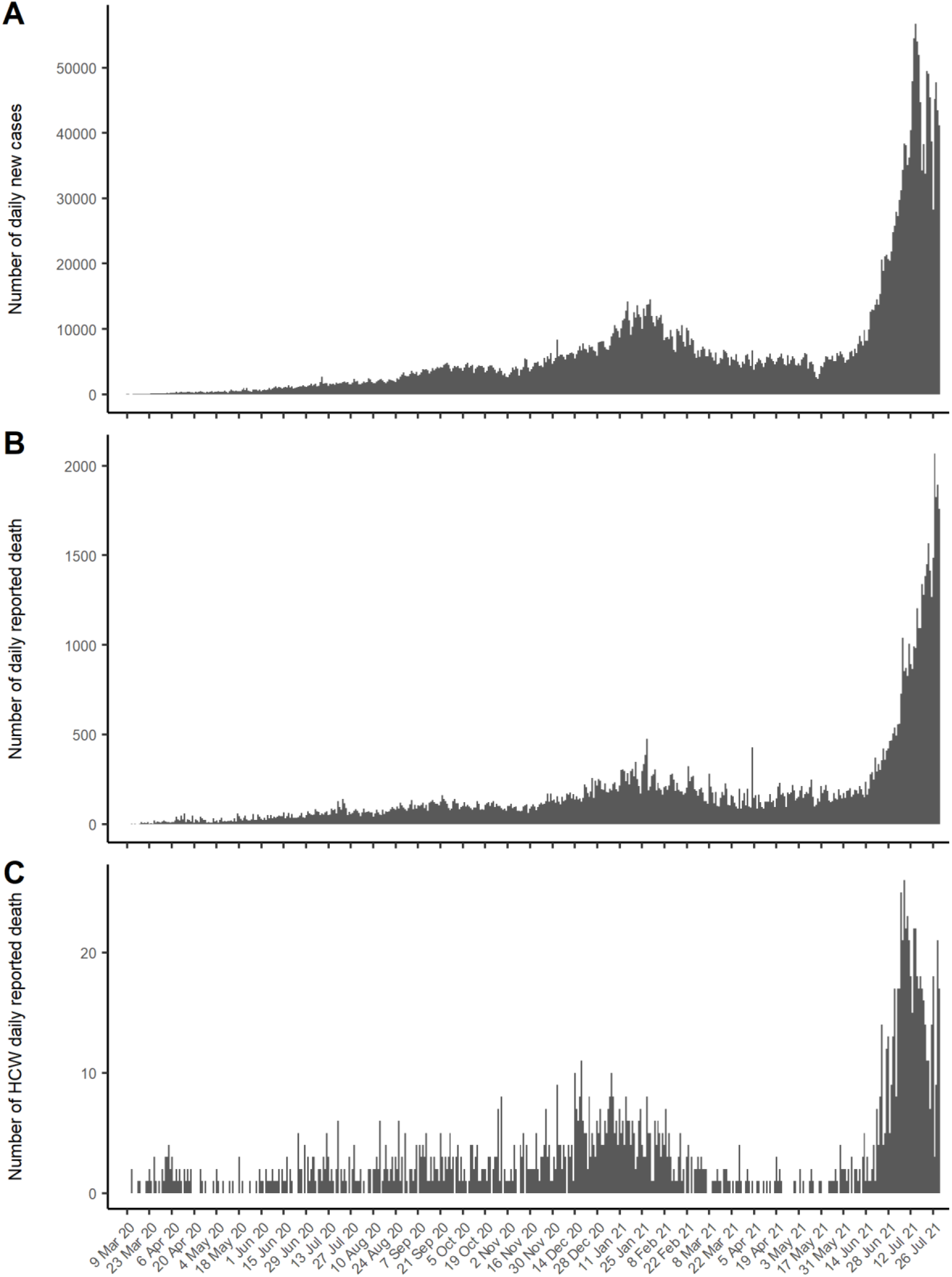
Daily reported new cases (A), daily reported deaths (B) and daily reported HCW deaths between March 2020 and July 2021 in Indonesia.

Vaccination campaign for health care workers initiated since mid of January 2021. A month later, the MoH claimed that 85% of targeted HCWs were vaccinated except those with comorbid or pregnant [43, 44]. *Pusara Digital* recorded HCW deaths prior to the launch of the vaccination program accounting for over a third of total HCW deaths (Table 1). We were only able to obtain the vaccination status of 116 (7.5%) HCWs because vaccination status for nearly half of the deceased was not available publicly. Among the available data, 54 (12%) involved with COVID-19 patient care and had an underlying health condition.

### Mortality rate of HCW population and the general population

The number of daily new infections first peaked at just under 15,000 during early 2021 (Figure 1A), then steadily declined and leveled at about 6,000 until June 2020. Thereafter, a very substantial surge occurred, peaking at 56,575 cases on 15 July 2021. COVID-19 caused a 10-fold surge in mortality attributable to COVID-19 (2069 deaths on 27 July 2021 vs. typically <200 daily since early 2020, Figure 1B).

Figure 1C implies the mean of mortality rate in the first six months of the COVID-19 pandemic reached 0.04/1000 HCW and accounted for 0.11/1000 HCW in the second six months (September 2020 – February 2021). Between March and July 2021, the mean of crude mortality rate was 0.16/1000 HCW. Mortality rate in July 2021 was higher than crude mortality rate from the previous months (0.56/1000 vs. 0.07/1000).

Table 2 shows the COVID-19 mortality rate of the HCW population at the national, and provincial level. Between March 2020 and July 2021, the national HCW mortality rate was 1.707 deaths per 1000 HCWs. The HCW mortality rate on Java was 2.883 deaths per 1000 HCWs, significantly higher than the national HCW mortality rate (p< 0.001). About 46% of HCWs and 60% of the Indonesian population live on Java Island, and account for 71% of national COVID-19 total deaths.

**Table 2.** Mortality rates of HCW and general population in Indonesia between March 2020 and July 2021.

| Province/Island | Health workers |  |  | General Population<sup>3</sup> |  |  | Death Relative Ratio |  |
| --- | --- | --- | --- | --- | --- | --- | --- | --- |
|  | Population<sup>1</sup> | Reported death<sup>2</sup> | Mortality rate (per 1000) | Population | Reported death | Mortality rate (per 1000) | Ratio | 95% CI |
| **Sumatera** |  |  |  |  |  |  |  |  |
| Aceh | 40,110 | 18 | 0.449 | 5,459,891 | 988 | 0.181 | **2.48** | **1.56 - 3.95** |
| North Sumatera | 48,841 | 53 | 1.085 | 14,703,532 | 1,465 | 0.100 | **10.89** | **8.28 - 14.32** |
| West Sumatera | 22,416 | 9 | 0.401 | 5,498,751 | 1,496 | 0.272 | 1.48 | 0.77 - 2.84 |
| Riau | 20,717 | 24 | 1.158 | 7,128,305 | 2,592 | 0.364 | **3.19** | **2.13 - 4.76** |
| Jambi | 17,636 | 7 | 0.397 | 3,677,894 | 425 | 0.116 | **3.43** | **1.63 - 7.25** |
| South Sumatera | 32,666 | 15 | 0.459 | 8,567,923 | 2,055 | 0.240 | **1.91** | **1.15 - 3.18** |
| Bengkulu | 11,040 | 5 | 0.453 | 2,019,848 | 285 | 0.141 | **3.21** | **1.33 - 7.77** |
| Lampung | 23,912 | 24 | 1.004 | 8,521,201 | 2,006 | 0.235 | **4.26** | **2.85 - 6.37** |
| Bangka Belitung | 6,692 | 1 | 0.149 | 1,517,590 | 663 | 0.437 | 0.34 | 0.05 - 2.43 |
| Riau Islands | 8,086 | 7 | 0.866 | 2,242,198 | 1,161 | 0.518 | 1.67 | 0.80 - 3.51 |
| **Total** | **232,116** | **163** | **0.702** | **59,337,133** | **13,136** | **0.221** | **3.17** | **2.72 - 3.70** |
| **Java** |  |  |  |  |  |  |  |  |
| DKI Jakarta | 57,439 | 180 | 3.134 | 10,644,986 | 12,173 | 1.144 | **2.74** | **2.37 - 3.17** |
| West Java | 106,128 | 213 | 2.007 | 49,935,858 | 9,363 | 0.188 | **10.70** | **9.35 - 12.26** |
| Central Java | 105,626 | 187 | 1.770 | 34,940,078 | 19,343 | 0.554 | **3.20** | **2.77 - 3.69** |
| DI Yogyakarta | 17,800 | 19 | 1.067 | 3,882,288 | 3,401 | 0.876 | 1.22 | 0.78 - 1.91 |
| East Java | 106,210 | 565 | 5.320 | 39,886,288 | 20,330 | 0.510 | **10.44** | **9.60 - 11.34** |
| Banten | 25,767 | 44 | 1.708 | 13,160,496 | 1,923 | 0.146 | **11.69** | **8.67 - 15.75** |
| **Total** | **418,970** | **1,208** | **2.883** | **152,449,994** | **66,533** | **0.436** | **6.61** | **6.24 - 6.99** |
| **Bali and Nusa Tenggara** |  |  |  |  |  |  |  |  |
| Bali | 20,097 | 13 | 0.647 | 4,380,824 | 2,151 | 0.491 | 1.32 | 0.76 - 2.27 |
| West Nusa Tenggara | 18,458 | 11 | 0.596 | 5,125,622 | 568 | 0.111 | **5.38** | **2.96 - 9.76** |
| East Nusa Tenggara | 20,652 | 5 | 0.242 | 5,541,394 | 725 | 0.131 | 1.85 | 0.77 - 4.46 |
| **Total** | **59,207** | **29** | **0.490** | **15,047,840** | **3,444** | **0.229** | **2.14** | **1.49 - 3.08** |
| **Kalimantan** |  |  |  |  |  |  |  |  |
| West Kalimantan | 16,780 | 9 | 0.536 | 5,134,760 | 650 | 0.127 | **4.24** | **2.20 - 8.18** |
| Central Kalimantan | 13,036 | 9 | 0.690 | 2,769,156 | 861 | 0.311 | **2.22** | **1.15 - 4.28** |
| South Kalimantan | 17,283 | 22 | 1.273 | 4,303,979 | 1,351 | 0.314 | **4.06** | **2.53 - 6.17** |
| East Kalimantan | 14,580 | 27 | 1.852 | 3,793,152 | 3,345 | 0.882 | **2.10** | **1.44 - 3.07** |
| North Kalimantan | 4,099 | 1 | 0.244 | 768,505 | 348 | 0.453 | 0.54 | 0.08 - 3.83 |
| **Total** | **65,778** | **68** | **1.034** | **16,769,552** | **6,555** | **0.391** | **2.64** | **2.08 - 3.36** |
| **Sulawesi** |  |  |  |  |  |  |  |  |
| North Sulawesi | 11,867 | 5 | 0.421 | 2,528,794 | 704 | 0.278 | 1.51 | 0.63 - 3.65 |
| Central Sulawesi | 16,177 | 5 | 0.309 | 3,096,976 | 642 | 0.207 | 1.49 | 0.62 - 3.59 |
| South Sulawesi | 40,256 | 48 | 1.192 | 8,928,004 | 1,338 | 0.150 | **7.96** | **5.97 - 10.61** |
| Southeast Sulawesi | 15,857 | 3 | 0.189 | 2,755,589 | 356 | 0.129 | 1.46 | 0.47 - 4.56 |
| Gorontalo | 5,456 | 1 | 0.183 | 1,219,576 | 228 | 0.187 | 0.98 | 0.14 - 6.99 |
| West Sulawesi | 7,379 | 1 | 0.136 | 1,405,012 | 170 | 0.121 | 1.12 | 0.16 - 8.00 |
| **Total** | **96,992** | **63** | **0.650** | **19,933,951** | **3,438** | **0.172** | **3.77** | **2.94 - 4.83** |
| **Maluku and Papua** |  |  |  |  |  |  |  |  |
| Maluku | 8,349 | 4 | 0.479 | 1,831,880 | 226 | 0.123 | **3.88** | **1.45 - 10.43** |
| North Maluku | 6,693 | 2 | 0.299 | 1,278,764 | 238 | 0.186 | 1.61 | 0.40 - 6.46 |
| West Papua | 5,593 | 4 | 0.715 | 981,822 | 285 | 0.290 | 2.46 | 0.92 - 6.61 |
| Papua | 11,563 | 4 | 0.346 | 3,435,430 | 264 | 0.077 | **4.50** | **1.68 - 12.08** |
| **Total** | **32,198** | **14** | **0.435** | **7,527,896** | **1,013** | **0.135** | **3.23** | **1.91 - 5.47** |
| **Indonesia** | **905,261** | **1,545** | **1.707** | **271,066,366** | **94,119** | **0.347** | **4.92** | **4.67 - 5.17** |

Data sources:
<sup>1</sup>The number of HCWs (medical doctors, nurse, midwives, pharmacists, laboratory technicians, public health staff) and the number of population at provincial level was extracted from the 2020 Annual Indonesia Health Profile by the Ministry of Health
<sup>2</sup>The number of HCWs death was extracted from the *Pusara Digital*, https://nakes.laporcovid19.org/
<sup>3</sup>The number of general population was extracted from the Emerging Infectious Disease website, Ministry of Health, https://infeksiemerging.kemkes.go.id/dashboard/covid-19

Figure 2A illustrates varied provincial level of HCW mortality rates, ranging from 0.136/1000 (West Sulawesi) to 5.32/1000 (East Java). There were five provinces where HCW mortality rates were higher than the national rate: Central Java (1.770); East Kalimantan (1.851); West Java (2.001); Jakarta (3.134); and East Java (5.319).

**Figure 2.**
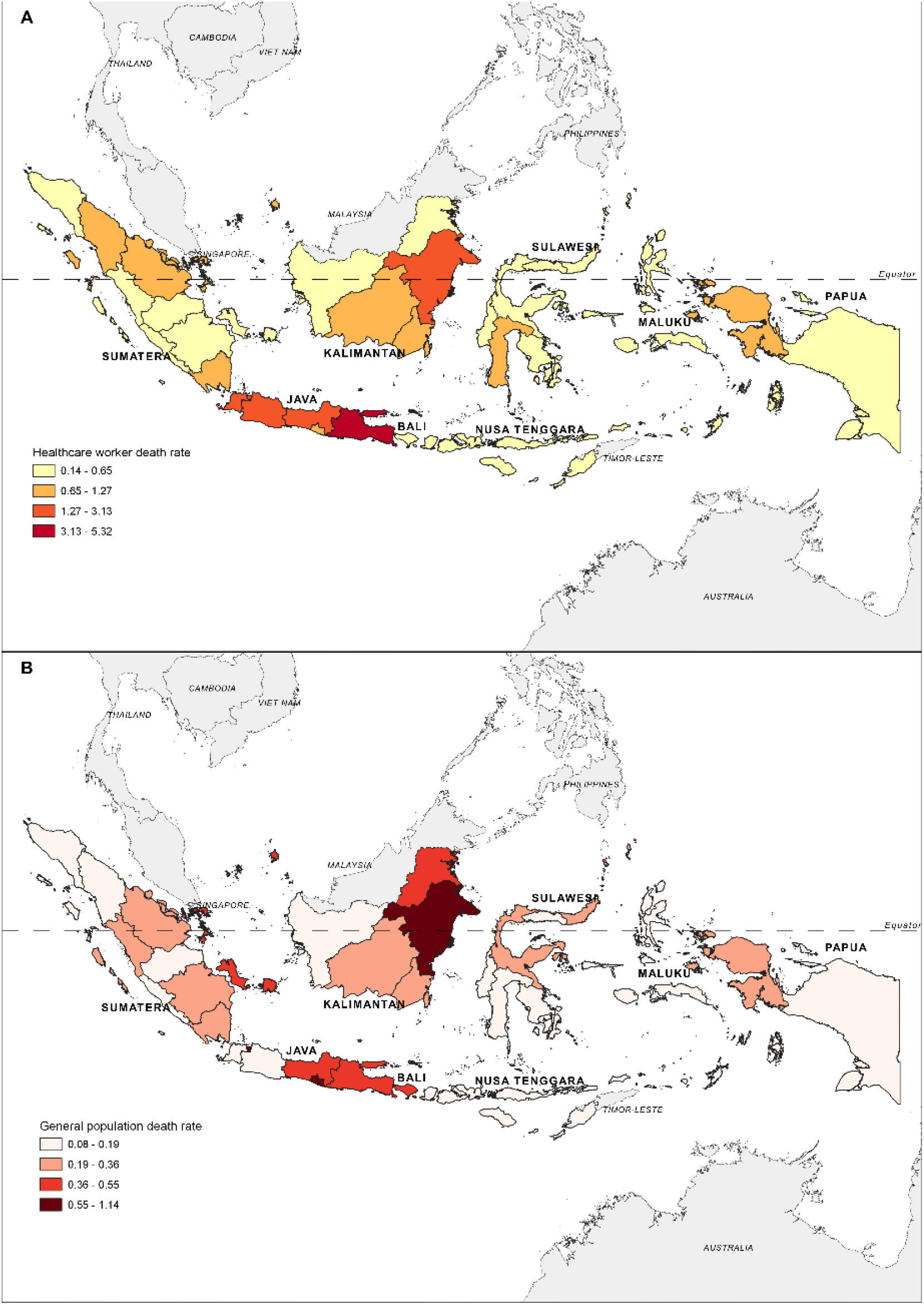
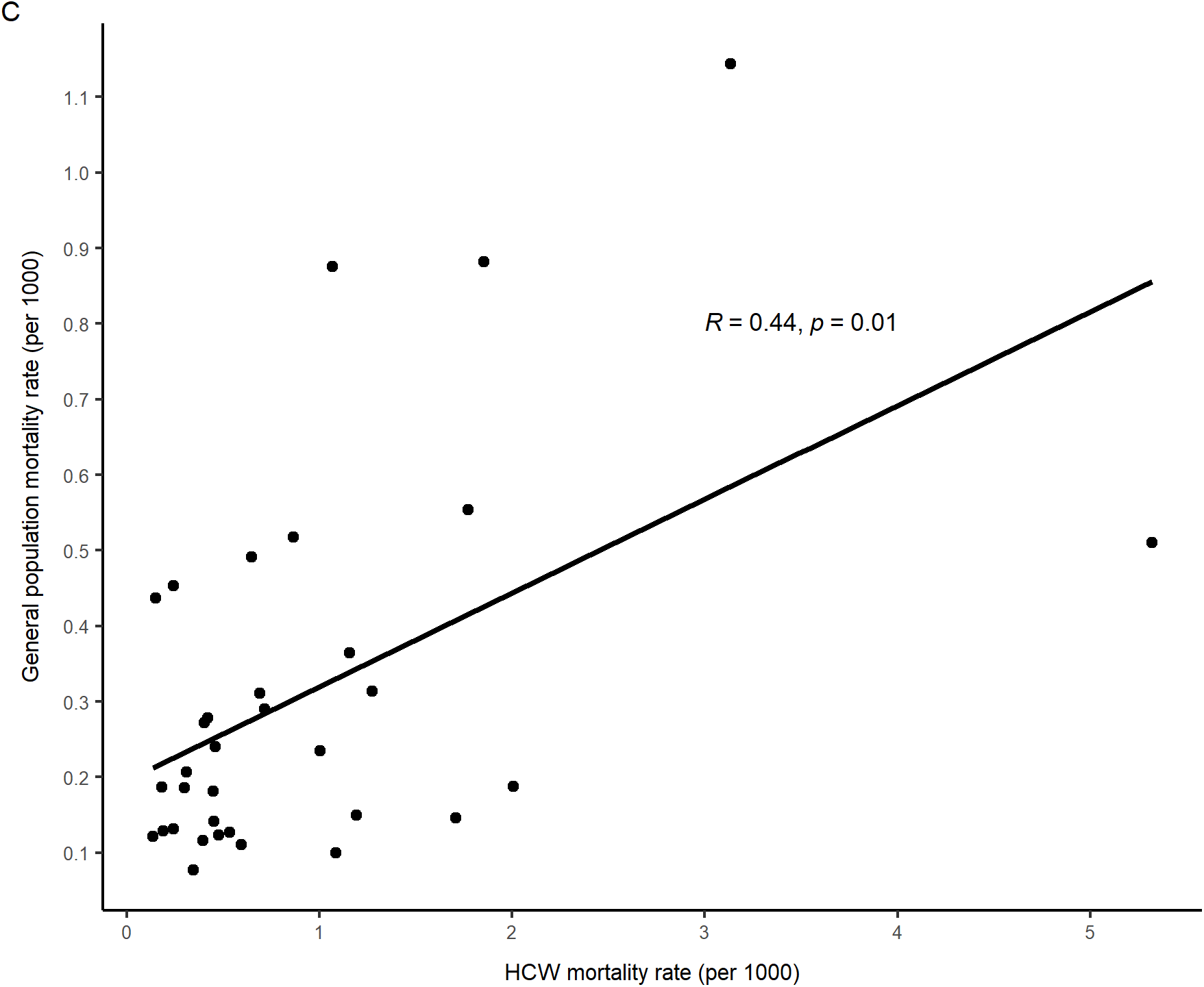
COVID-19 mortality rates on healthcare workers (A), general population (B) and the scatter plot between mortality rate for healthcare workers and general population during March 2020 and July 2021 in Indonesia.

The COVID-19 mortality rate of the general population was 0.347 deaths per 1000 people during the same period. Figure 2B illustrates that rate at the provincial level ranging from 0.077/1000 people (Papua) to 1.144/1000 people (Jakarta). Among provinces, the COVID-19 mortality rate among HCW correlated with that in the general population (p = 0.009) (Figure 2C).

Table 2 illustrates the death risk ratio between HCWs and the general population. The national death risk ratio of HCW against the general population was 4.92 (95% CI 4.67 – 5.17). In Java, the death risk ratio of HCW was significantly higher than the general population (RR = 6.61, 95% CI 6.24 – 6.99). Provincial death risk ratios ranged from 0.34 (95% CI 0.05 – 2.43; Bangka Belitung Islands) to 11.69 (95% CI 8.67 – 15.75; Banten).

### Community Responses on HCWs Mortality

We performed content analysis to understand messages in the submitted testimonials, identify patterns in the set of texts and explore their concerns about the loss. By the end of July 2021, *Pusara Digital* received 2,606 testimonials from colleagues (1,377), family, and friends (393), community (362), students-teachers (357), and former patients (118) of the deceased.

The coding process identified three major themes from the testimonials: revealing the valued personality of the deceased (5,196 comments), providing comfort and compassion to the family and HCW on duty (2,134 comments), and wishing peaceful afterlife (1,664 comments). By considering the complexity of the testimonials and in order to understand the content, multiple codes were allowed.

The senders recalled that the late HCWs possessed valued personalities (5,196 comments). Most comments identified them as loving and care individuals (664 comments), noticeably happy and optimistic persons (547 comments), dedicated (321 comments) and professional (315 comments), low profile (307 comments), treat others with respect (307 comments), and attentive (275 comments).

> *“Following his departure, I read a lot of positive impressions and stories about him. He was kind, friendly and helpful with an unforgettable gentle smile. His advice was always conveyed in simple words yet full of meaning. Deep condolences came not only from the family, but also from his students and colleagues. Our role model has gone but the memory of him stays forever”* (Niece of the deceased). Fond memories associated with personal life, heroic and/or professional experiences of the late HCWs also appeared among the testimonials: *“She was a nurse in our pediatric ward before transferring to the [COVID-19] care unit. Always put the patients as her high priority, loyal and dedicated. Her last wish if she did not make it [due to COVID-19 infection] was being buried in her honored uniform”* (Senior colleague of the deceased).

The second most frequent testimonials provide comfort and compassion. The participants passed farewell messages (657 comments) and deep condolences (472 comments), along with gratitude and acknowledgement of their service (457 comments), and support for the struggling family (176 comments). One of them is as the following:

> *“The deceased acted as my aunt, and senior colleague. Her story while working in a remote and impoverished areas in the eastern part of Indonesia was inspired me to become a doctor. Thank you for nurturing me although you did not realize it. I deeply lost a generous and low-profile figure. Happy reunited with your husband in heaven”* (Family of the deceased).

Furthermore, the community and health facility users (104 comments) also provided support to the left-behind family. They encouraged the HCWs on duty to save more lives and continue the fights against pandemic.

> *“May God grant heaven to the HCWs who died during this pandemic. May the left-behind family be strong and encouraged as their loved ones died in carrying out a noble task to save as many lives as possible. Keep fighting and keep up the spirit in the air, front-liners. God will always protect all of you”* (Community member).

*Pusara Digital* recorded 1,000 comments praying for peaceful life after death and rewards for their good deeds (664 comments). The death of HCWs brought pains not only for the family (433 comments), but also for their colleagues (557 comments), and teacher/students (394 comments).

> *“Despite the painful separation, I am proud of my father who consistently served people until the last days of his life. As a health worker, his noble fight was not in vain and will be eternally remembered. Rest in peace, Dad”* (Son of the deceased).

## Discussion

This first report shows the devastating impact of the COVID-19 pandemic to the Indonesian health workforce. We found at least 1,545 HCWs were lost during the early COVID-19 waves of 2020 and 2021. The national HCW mortality rate was thus estimated at 1.707/1000. There is no record of confirmed case numbers among HCW per se. Indonesian professional healthcare organizations estimated at least 15,000 of their members had been infected by the SARS-CoV-2 virus during 18 months of pandemics [45].

A crude case fatality rate for HCWs may thus be estimated at 10.3%, or 3-times higher than the general population. If true, this points to some features of hospital exposure as exacerbating the risk of death, e.g., higher infectious dose. The rapid loss of HCW is alarming due to relatively low numbers of health practitioners, Indonesia’s health services were stretched even before COVID-19, with only two doctors and 15 nurses per 10,000 population in 20 [30]. In comparison, people in the Philippines are served by six doctors and 49 nurses [46], while Malaysia is assisted by 19 doctors and 34 nurses [47]. This study suggests that the death rate of Indonesian HCWs was higher than United States (0.49 deaths per 1000 HCWs), Brazil (0.46) and India (0.25), but lower than in Peru (8.48) and Mexico (3.8) [48-52].

Observational retrospective studies are required to calculate an estimate of infections and excess deaths of HCWs attributable to COVID-19. However, a credible estimate of HCWs mortality amounted to all SARS-CoV-2 infections was unavailable in Indonesia. A study reported that among 1201 specimens collected from HCWs in early pandemic in Jakarta Province, 7.9% were confirmed positive for SARS-CoV-2 with the majority coming from medical doctors and nurses (44.2%) [53]. Most of them (64%) were reported having in (close?) contact with suspect/confirmed COVID-19 cases. The screening rate among HCWs remains low in early pandemic [53]. The number of reported deaths may be lower than the actual deaths due to COVID-19. Indonesia only counted and reported deaths with molecular confirmation and neglected counting deaths with no molecular testing (probable death). The issue may come from variable capacities of testing and tracking among provinces.

Investment in the national surveillance system and sharing data of infections and deaths are important to enable preventive actions. Similar issues have occurred in the United States, Italy, and the United Kingdom, where data were limited and challenging to obtain [54]. Essential data on socio-demographic, occupational type, working place, and vaccination status remained difficult to collect from official sources. As an alternative, we engaged with the journalist’s network and volunteers to fill the gap by curating from social media as well as contacting medical and healthcare organizations.

Previous studies revealed that proper grief responses and mourning process had reduced mental health burden among the devastated families and colleagues [55, 56]. However, strict implementation of physical distancing obviously forced families from holding proper funerals and cultural death ceremonies [55]. *Pusara Digital* serves as an alternative service to the conventional ritual of grave pilgrimage, particularly for those who were unable to attend the funeral [57]. The platform offers post-funeral activities, including commemoration, care and support for families and friends.

Moreover, financial compensation for the deceased’s family is essential to support the livelihood of families. The Indonesian government regulated legal basis of financial compensation for those HCWs who provided care for COVID-19 patients in hospitals and/or died due to COVID-19 infections [58]. By end of July 2021, only 218 families of deceased HCWs confirmed receiving the financial compensation. Thus, community groups and medical and healthcare associations worked on advocacy and reimbursement to ensure all families receive it [59].

Our study has limitations within which our findings need to be interpreted carefully. First, the individual data were obtained from *Pusara Digital* records. There were 35% of missing age and approximately 48% of vaccination status were inaccessible. Second, age and gender distribution of HCWs in Indonesia are publicly unavailable. Consequently, we were unable to calculate standardized age and gender mortality rate and improve the interpretation of the relative risk compared to the general population at risk from COVID-19. Last but not least, content analysis emphasizes on words and involves subjective interpretation, which may diminish the context and affect the reliability of the results.

Finally, this study suggests that the COVID-19 pandemic confirms the high priority need of strong healthcare systems against any emerging pandemic threats. Further transformation in advanced molecular diagnostic research and technology, rapid community surveillance and epidemiological and clinical studies, integrated data management, data transparency and data analysis, information technology for supply chain, trainings, communication, workforce management is crucial to protect and response next epidemic.

## Conclusion

This study finds that the COVID-19 event in Indonesia resulted in the loss of many hundreds of HCW, most of them being senior physicians, nurses, and midwives. They died at rates 5-times higher than everyone else. The sheer sparseness of the workforce requires more protective steps and a national systematic surveillance of occupational mortality is urgently needed in this setting.

## Data Availability

All data produced in the present study are available upon reasonable request to the authors
All data produced in the present work are contained in the manuscript
All data produced are available online at Pusara Digital; https://nakes.laporcovid19.org/ and the Ministry of Health in the Republic of Indonesia. https://infeksiemergingkemkesgoid/dashboard/covid-19. 2022.

## Acknowledgement

We express deepest condolences and sincere gratitude to the family, friends and colleagues of Indonesian Healthcare Workers who lost their lives during the COVID-19 pandemic. We also thank all participants who have provided testimonials to commemorate their life.

The authors indebt sincere gratitude to the following dedicated volunteers in *Pusara Digital* team: Iskandar Muhad, Sonny Prayogo, Fakhriy Fathur, Cici Riesmasari, Jeremy H. Limbong, Hilman Arioaji, Maria Bramanwidyantari, Indah Kartika Cahyani, Yoesrianto, Jeanne Eureka, Fransiska R. Simbolon, Alfia Nadia Putri, Lutfiyyah Haniffariyani, Dwikita Ichsana, Yoesep Budianto, and Windyah Puji Lestari.

Pusara Digital, the main source of this study, was partially funded by the Digital Access Program of the British Embassy in Indonesia and as part of collaborative work with the Association of Indonesian Medical Doctors (*Ikatan Dokter Indonesia; IDI*), National Association of Nurses (*Persatuan Perawat Nasional Indonesia, PPNI*), Association of Midwives (*Ikatan Bidan Indonesia, IBI*), Association of Medical Laboratory Technologists (*Persatuan Ahli Teknologi Laboratorium Kesehatan Indonesia, PATELKI*), and Eijkman-Oxford Clinical Research Unit (EOCRU).

## Author Contributions

ICMJE criteria for authorship read and met: LLE, AA, IH, AN, MZM, KDL, AT, FF, FN, QESA, HM, TM, AR, MP, KA, AAA, NN, AVSS, AJ, NSW, HS, HWS, ADS, PK, JKB, IRFE. Agree with the manuscript’s results and conclusions: LLE, AA, IH, AN, MZM, KDL, AT, FF, FN, QESA, HM, TM, AR, MP, KA, AAA, NN, AVSS, AJ, NSW, HS, HWS, ADS, PK, JKB, IRFE. Designed the experiments/the study: LLE, IRFE. Analyzed the data: LLE, AA, MZM, KDL, IRFE. Collected data/did experiments for the study: LLE, AA, MZM, KDL, AT, FF, FN, QESA, HM, TM, AR, MP, KA, AAA, NN, AVSS, AJ, NSW. Wrote the first draft of the paper: LLE, IRFE. Contributed to the writing of the paper: LLE, AA, IH, AN, KDL, QESA, AH, HWS, ADS, PK, JKB, IRFE.

